# Interoceptive accuracy and attention across multimorbidity classes: A latent class analysis

**DOI:** 10.64898/2026.06.08.26355147

**Authors:** Jesper Mulder, C. Madeleine Böker, Anne K. Smit, Jessica C. Kiefte-de Jong

**Author notes:** Contact information corresponding author: Jesper Mulder; Health Campus the Hague, Department of Public Health and Primary Care, Leiden University Medical Center, The Hague, The Netherlands; Turfmarkt 99, 2511 DP The Hague, The Netherlands.

## Abstract

**Background:** Multimorbidity is increasingly prevalent, and associated with worse clinical and psychosocial burdens. Interoception, the brain’s ability to sense and interpret internal bodily signals, may contribute to multimorbidity, through its link with health behaviors, stress regulation, and mental health. This study examines whether self-reported interoceptive accuracy and attention is associated with multimorbidity, by identifying multimorbid subgroups and their interoceptive profiles.

**Methods:** Morbidity classes were identified through latent class analyses in two Dutch survey datasets, focusing on depression and alexithymia (DA-dataset; N = 671) and lifestyle factors (L-dataset; N = 1022). Linear regression analyses were used to assess interoceptive accuracy and attention (by the Interoceptive Accuracy Scale and Interoceptive Attention Scale respectively) among different subgroups.

**Results:** Multimorbid subgroups were characterized by older age, low socioeconomic position, and elevated physical, psychological, and behavioral problems. Multimorbid classes exhibited lower interoceptive accuracy (DA-dataset: B = −1.14, 95% CI = [−2.89, 0.62]; L-dataset: B = −2.36, 95% CI = [−3.83, −0.89]) and higher attention (DA-dataset: B = 3.62, 95% CI = [0.97, 6.27]; L-dataset: B = 1.07, 95% CI = [−1.42, 3.56]) compared to healthier classes.

**Conclusion:** Multimorbid populations demonstrated lower interoceptive accuracy and higher interoceptive attention. This highlights the psychosocial complexity of multimorbid populations which may impact their self-management and health behavior. These findings underscore the need to expand treatments to include psychosocial domains for multimorbid patients.

**Highlights:**

- Interoceptive dysregulation and multimorbidity share risk factors
- Multimorbid populations show low interoceptive accuracy but high attention
- Multimorbid populations have distinct interoceptive profiles and psychosocial risks
- Embedding interoception in multimorbid care may help personalize treatments

## 1. Introduction

The prevalence of multimorbidity (i.e., the presence of at least two coexisting chronic conditions without a clear hierarchy; van den Akker et al., 1996) is steadily increasing, particularly among older adults (Chen et al., 2020; Souza et al., 2021). Multimorbidity is associated with adverse clinical outcomes (e.g., higher mortality and restricted physical functioning) as well as psychosocial burdens (e.g., increased risk of mental health issues, reduced quality of life, and greater difficulties in self-management; Bayliss et al., 2007; Chen et al., 2020) compared to individuals with single or no chronic conditions. The health situation of multimorbid patients is complex, because often both physical and psychological conditions are present in these populations. For instance, among multimorbid patients depression is two to three times more prevalent compared to those with one or no chronic condition (Read et al., 2017). Additionally, multimorbidity is associated with a higher prevalence of unhealthy lifestyle behaviors, such as low physical activity (Delpino et al., 2022), sleep problems (Zhou et al., 2023), smoking, and alcohol use (Alvarez-Galvez et al., 2023; Barnett et al., 2012; Bayliss et al., 2007; Larsen et al., 2017). This co-occurrence of somatic, psychological, and behavioral factors challenges the current healthcare system.

Psychosocial problems may affect interoception (i.e., the brain’s ability to sense and interpret internal bodily signals), and disturbed interoception may influence psychosocial problems. For example, symptoms of depression have been linked to reduced cardiac interoception, and not being able to appropriately anticipate interoceptive states has been linked to increased risk of anxiety disorders (Khalsa et al., 2018; Quadt et al., 2018). Interoception can be subdivided into interoceptive accuracy, referring to whether individuals are accurate in perceiving internal signals, and interoceptive attention, which reflects the extent to which individuals pay attention to these signals (Murphy et al., 2019). Interoception plays a critical role in shaping health-related outcomes by influencing homeostasis (i.e., the regulation of internal physiological balance) through autonomic reflexes, consciously perceived internal signals and bodily needs, and modulating sickness behaviors such as fatigue, anhedonia, and social withdrawal (Farb et al., 2015; Quadt et al., 2018) and self-management (Guo et al., 2026). For instance, when homeostatic balance is threatened, such as during energy depletion, a hunger-related interoceptive signal prompts the behavioral response needed to restore it (Farb et al., 2015). As a result, interoception and health could be bidirectionally linked: on the one hand interoception could influence health behaviors that influence cardiovascular and metabolic disease risk (Mulder et al., 2025). On the other hand, several cardiometabolic diseases (Bonaz et al., 2021), alexithymia (i.e., difficulty being aware of and identifying feelings) and depression (Eggart et al., 2019; Shah et al., 2016), and alcohol use disorders (Wisniewski et al., 2021) in itself come along with disrupted interoception. When interoception becomes maladaptive, characterized by excessive or insufficient attention to bodily cues and systematic misinterpretation (Trevisan et al., 2023), individuals may experience poor self-regulation, and worsening mental and physical health (Farb et al., 2015; Weiss et al., 2014). Such a pattern may be particularly important in the context of multiple co-occurring conditions, where accurate interpretation of bodily signals is essential for recognizing symptoms, adhering to treatment, and adjusting health behaviors across interacting diseases.

Given that interoceptive dysregulation and multimorbidity appear to share common risk factors and may mutually reinforce one another, examining their relationship could reveal important mechanisms underlying disease clustering. Interoceptive dysregulations frequently occur within vulnerable populations that are already at heightened risk for multimorbidity, such as older adults, individuals with low socioeconomic position, and those with elevated stress, poor sleep, increased body mass index, or unhealthy lifestyle behaviors such as excessive alcohol intake and unhealthy dieting (Davidson & Stevenson, 2022; Ewing et al., 2017; Murphy et al., 2018; Schulz & Vogele, 2015; Wisniewski et al., 2021). These patterns suggest that maladaptive interoception and multimorbidity frequently converge in the same high-risk populations. However, little is known about how interoceptive processes relate to multimorbidity patterns, or whether interoceptive accuracy and attention associate differently with distinct disease clusters. Therefore, the current study aims to address these gaps by examining how self-reported interoceptive accuracy and attention relate to multimorbidity in Dutch adults. Specifically, we aimed to 1) identify subpopulations in Dutch adults experiencing multimorbidity and explore the psychological, social and behavioral conditions that co-occur with this clustering of diseases, and 2) examine the relationship between self-reported interoceptive accuracy and attention and identified subpopulations with varying disease statuses and psychosocial backgrounds.

## 2. Methods

### 2.1. Study Design

Data collection was conducted through research organization Flycatcher using a cross-sectional survey design between July 2024 and August 2024. Flycatcher is an independent Dutch online panel of over 10,000 adults, designed to give a broad representation with respect to age, sex, region, and socio-economic position (Flycatcher, 2026). Participants were at least 18 years old, Dutch speaking, and enrolled through a double-active opt-in mechanism. The panel remained active until the minimum required sample size was reached. Incentives in the form of points were gifted for participation, which could be exchanged for vouchers. The local research committee (reference number WSC-2024-26/ SP) and local ethics committee (METC number N24.072) approved this study as non-WMO. Flycatcher adheres to ISO 20252 and ISO 27001 quality standards.

Data collection was part of a larger research project and resulted in two separate datasets: one included variables on depression and alexithymia (“DA-dataset”), and the second included lifestyle factors (“L-dataset”). Sample size calculations were performed for both datasets separately. The DA-dataset sample size calculation was based on an effect size of 0.2, an alpha level of 0.05, a power of 0.8, and four predictors, resulting in a sample size of 815. The L-dataset used the same effect size, alpha, and power, but with ten predictors, resulting in a sample size of 1240. Both samples exceeded the minimum of 300 participants recommended for latent class analysis (Weller et al., 2020).

### 2.2. Material

All questionnaires were administered in Dutch. The included questionnaires, along with their corresponding items, response scales, and internal consistencies are presented in Table 1. All questionnaires showed good to excellent internal consistency (α = 0.72-0.96; ω = 0.81-0.96) in the current sample, except for the Alcohol Use Disorders Identification Test – Consumption (AUDIT-C), which showed unacceptable to questionable internal consistency (α = 0.45; ω = 0.62). The unacceptable Cronbach’s alpha and its discrepancy with McDonald’s omega is expected for the AUDIT-C, since the three items of the scale (i.e., frequency of drinking, typical quantity per occasion, and frequency of heavy drinking) are related but distinct facets of alcohol use. We chose to retain the variable, as the AUDIT-C was entered in the analyses as a validated binary categorical variable using validated cutoff scores (Duffy et al., 2023; Ingesson-Hammarberg et al., 2024; van Gils et al., 2021).

**Table 1.**
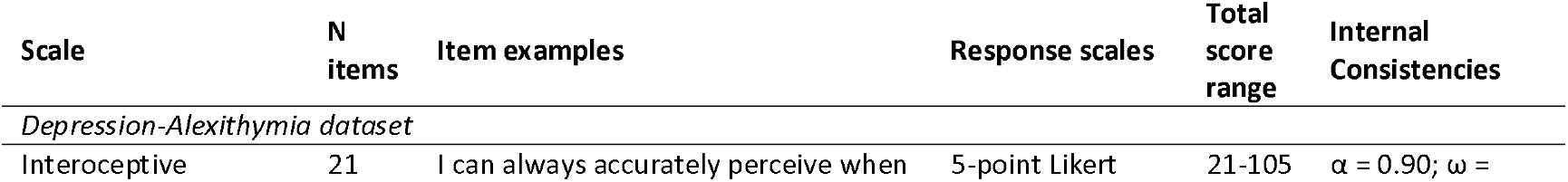
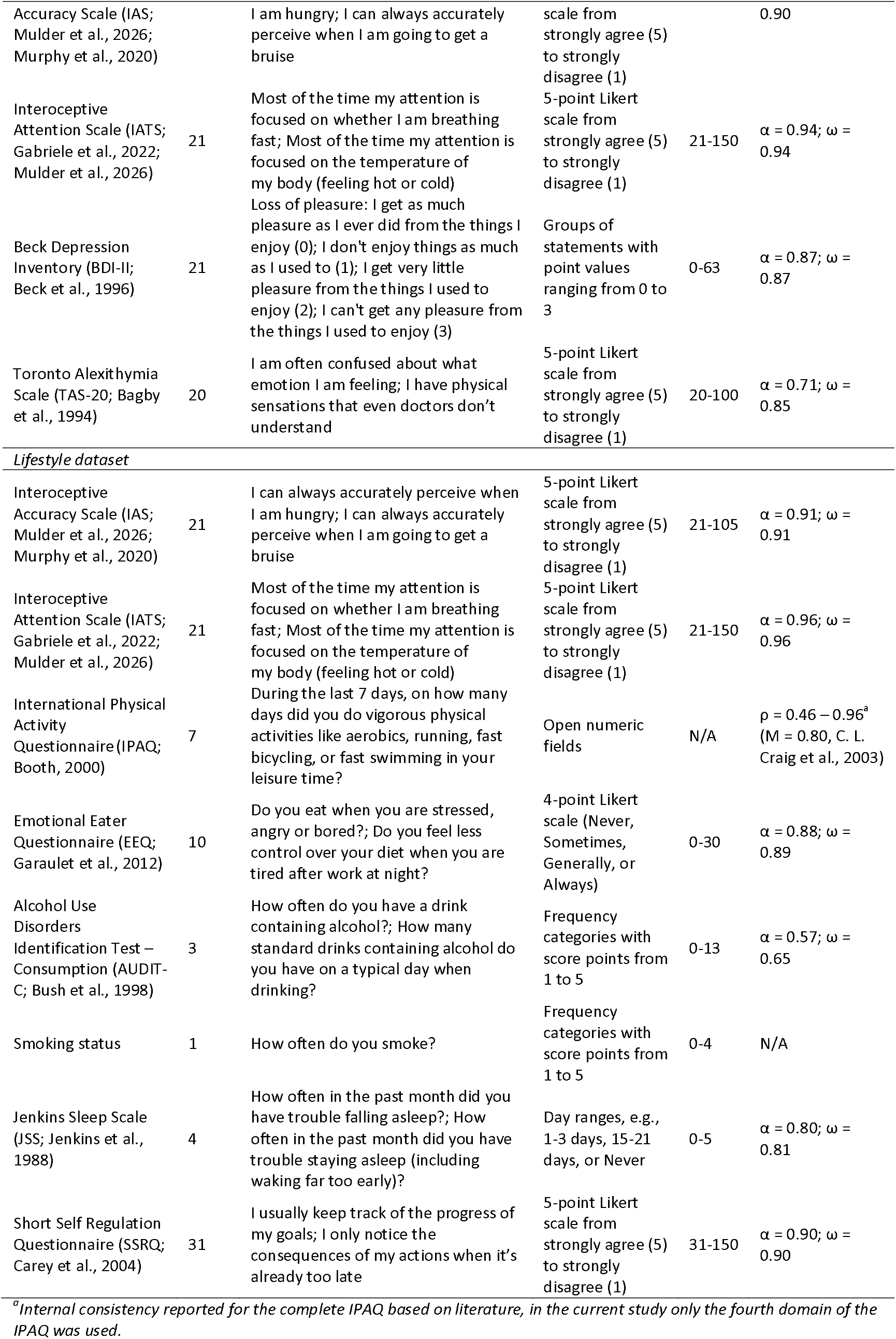
Questionnaires with their corresponding items, response scales, and current sample internal consistencies used during the data collection process between July 2024 and August 2024 in 671 (Depression-Alexithymia dataset) and 1022 (Lifestyle dataset) Dutch adults.

### 2.2.1. Categorization

The primary measures of interoceptive accuracy and attention remained continuous variables, whereas all other variables were categorized. Multimorbidity was assessed based on participants’ self-reported health status, with chronic conditions and risk factors separated from each other (Budreviciute et al., 2020). Chronic conditions included type 2 diabetes, asthma/COPD, cardiovascular diseases, cancer, muscle/joint problems, fatty liver disease, dementia, and other diseases. Disease Status was categorized as No Disease, One Disease, or Multimorbid based on the absence of, or the presence of one or more, of these chronic conditions. Risk factors included overweight or obesity, high blood pressure, and increased cholesterol. Risk factors were collapsed to a binary scale indicating whether these risk factors were absent or one or more of these risk factors were present.

Depression was categorized as Minimal depression (0-13), Mild depression (14-19), Moderate depression (20-28), or Severe depression (29-63) (Wang & Gorenstein, 2013). The TAS-20 scores were categorized as No alexithymia (0-60) or Alexithymia present (61-100) (Bagby et al., 1994).

Physical activity was derived from the leisure-time domain of the IPAQ, covering walking, moderate, and vigorous intensity activities. Weekly minutes were calculated per activity, and participants were classified as Insufficiently Active (<75 minutes vigorous or <150 minutes moderate intensity physical activity per week) or Physically Active (≥75 minutes vigorous or ≥150 minutes moderate exercise per week). Walking was considered moderate only if reported as moderate or fast paced. EEQ scores were divided into Non-emotional eater (0-5), Low emotional eater (6-10), Emotional eater (11-20), or Very emotional eater (21-30). AUDIT-C scores were categorized as no likelihood of alcohol dependency with scores below 4 for females and below 5 for males (Duffy et al., 2023; Ingesson-Hammarberg et al., 2024; van Gils et al., 2021). Smoking status was categorized as Non-Smoker or Smoker. JSS scores were split into No Disturbed Sleep (0-11) or Disturbed Sleep (12-20) (Juhola et al., 2021). SSRQ scores were divided by distribution-based percentiles into Below Average Self-Regulation or Above Average Self-Regulation.

Both datasets included the demographic variables age, sex, and socioeconomic position (SEP). Age was collapsed into binary levels, with the cut-off set at 45 years (with this age the prevalence for multimorbidity increases by 15,7% compared to 25–44-year-old individuals; Agborsangaya et al., 2012). Sex was measured binary. SEP was calculated based on self-reported education (low, middle, or high) and income (below average, average, or above average), and divided into Below average, Average, or Above average SEP (King et al., 2011).

### 2.3. Statistical analysis

#### 2.3.1. Data pre-processing

All data was inspected, cleaned, and analyzed using RStudio (version 2024.02) with R (version 4.3). Alpha was set at 0.05 a priori. Latent class analyses (LCA) required four assumption checks (Sinha et al., 2021). First, normal distributions of the continuous IAS and IATS outcome variables were checked based on skewness (values < |2| were assumed normal) and kurtosis (values < |7| were assumed normal) (Kim, 2013), and visually based on histograms and Q-Q plots. Second, multicollinearity was assessed using Cramer’s V. Cramer’s V greater than 0.5 indicated presence of multicollinearity. If variable pairs showed multicollinearity, they were assessed for importance to the central research question, measurement properties, and missingness, before a decision was made to drop or retain the variable(s). Third, frequency tables were examined for categories with less than 10% of the total sample. Categories that did not meet the threshold were collapsed. Finally, after modeling the LCA, local independence was checked. Within each latent class, observed predictors had to be independent of one another (residual correlations < 0.5; Sinha et al., 2021). If variable pairs violated the local independence assumption, they were assessed for importance to the research question, measurement properties, and missingness, before deciding to drop or retain the variable(s).

All continuous variables showed normal distribution. No variable pairs showed multicollinearity and local independence assumptions were not violated. Categories in the BDI-II and EEQ questionnaires included <10% of the total sample. Therefore, we collapsed these categories: BDI-II scores were dichotomized into “Low depressive symptoms” (0-13) and “Clinically relevant depressive symptoms” (>13) (Wang & Gorenstein, 2013); the EEQ “Very emotional eater” category was combined with the “Emotional eater” category into “Emotional to very emotional eater” (EEQ scores >10).

Missing values were removed listwise: in the DA-dataset 124 participants and in the L-dataset 192 participants had missing SEP components (education or income), causing SEP calculation to be impossible. Little’s MCAR test indicated missing values were consistent with missing at random. Outliers in the outcome variables were retained, as deviations may reflect meaningful variation in a clinical population. Extreme outliers based on z-scores > 3 and Mahalanobis distance critical values (chi-square, df = number of variables, p < .001) and impossible values were removed from the dataset for analysis. Impossible values included values that were outside the possible range of questionnaire individual items and total scores. Twenty outliers were removed from the DA-dataset, and four impossible values and 30 outliers were removed from the L-dataset.

#### 2.3.2. Data Analysis

In the descriptive analyses, continuous variables were presented as mean and standard deviation. Categorical variables were presented as frequencies and percentages.

A LCA was conducted using the package poLCA to identify subpopulations. Models with K = 1 to K = 10 classes were fitted, with 10 random starts per model. The number of latent classes was selected using a sequential decision procedure. First, models in which any class contained fewer than 10% of the sample were excluded. Second, the model with the lowest BIC was selected. When two or more models had equivalent BIC values, AIC was used as a tiebreaker. Entropy was reported as a class separation quality indicator, with values >.60 considered acceptable (Sorgente et al., 2025). Bootstrap Likelihood Ratio Tests (BLRT) were conducted for each sample, testing whether a more complex model showed better fit. BLRT served as supporting evidence for the BIC-based selection. To confirm stability, the selected model was re-estimated with 50 random starts. Convergence was checked by running 20 estimations and assessing replication of the maximum log-likelihood. Models were considered stable when all runs yielded log-likelihood values within 0.5% of the best-fitting solution and at least 20 runs replicated this value (Sinha et al., 2021). Models failing to meet these criteria were considered unstable and not interpreted.

To assess differences in self-reported interoceptive accuracy and attention, the classes identified in the LCA were used as predictors in regression analyses with IAS and IATS total scores as outcome variables. The class with the highest proportion of no chronic diseases was selected as the reference category, providing a clinically anchored baseline for class comparisons. If there were multiple classes with the same proportion of no chronic diseases, the lowest presence of risk factors was used as a tiebreaker.

Interoception in multimorbidity may be driven by underlying factors: aging may affect interoception (Li et al., 2025), there may be differences in interoception between sex (Prentice et al., 2022; Prentice & Murphy, 2022), and impaired interoception has been identified in depressed samples (Quadt et al., 2018). Therefore, regression analyses controlled for age (continuous), sex, and depression (BDI total scores, only in the DA-dataset) were performed as sensitivity analyses. Since these variables are defining features of the LCA classes, findings of the sensitivity analyses are a decomposition of why classes differ.

## 3. Results

### 3.1. Descriptives

The final sample consisted of 671 participants in the DA-dataset and 1022 participants in the L-dataset, after list-wise deletion of missing values and removal of impossible values. All descriptive statistics of both datasets are shown in Table 2.

**Table 2.** Descriptive statistics of 671 (DA-dataset) and 1022 (L-dataset) Dutch adults from the general population. Data was collected between July 2024 and August 2024.

|  | DA-dataset (N = 671) | L-dataset (N = 1022) |
| --- | --- | --- |
| **Continuous variables** | **M (SD)** | **M (SD)** |
| Age, years | 52.7 (16.5) | 52.1 (16.8) |
| Interceptive Accuracy Score (IAS) | 80.2 (9.8) | 81.4 (9.9) |
| Interceptive Attention Score (IATS) | 50.3 (14.9) | 57.2 (16.7) |
| **Categorical variables** | **N (%)** | **N (%)** |
| *Demographics* |  |  |
| Female sex | 318 (47.4) | 506 (49.5) |
| Age ≥45 years | 428 (63.8) | 639 (62.5) |
| SEP |  |  |
| Below average | 265 (39.5) | 342 (33.5) |
| Average | 119 (17.7) | 215 (21.0) |
| Above average | 287 (42.8) | 465 (45.5) |
| *Disease Status* |  |  |
| No chronic conditions | 403 (60.1) | 630 (61.6) |
| Single chronic condition | 182 (27.1) | 278 (27.2) |
| Multimorbidity | 86 (12.8) | 114 (11.2) |
| At least one risk factor present | 281 (41.9) | 423 (41.4) |
| *Psychological Variables* |  |  |
| Depression |  |  |
| Low depressive symptoms | 561 (83.6) |  |
| Clinically relevant depressive symptoms | 110 (16.4) |  |
| Alexithymia present | 153 (22.8) |  |
| *Lifestyle Behaviors* |  |  |
| Physically active |  | 586 (57.3) |
| Emotional eating |  |  |
| Non-emotional eater |  | 265 (25.9) |
| Low emotional eater |  | 331 (32.4) |
| Emotional to very emotional eater |  | 426 (41.7) |
| Likelihood of alcohol dependency |  | 136 (13.3) |
| Current smoker |  | 132 (12.9) |
| Disturbed sleep |  | 245 (24.0) |
| Below average self-regulation |  | 538 (52.6) |

### 3.2. Class Selection

Although the four-class model was algorithmically selected for the DA-dataset, this model indicated an unstable solution. Therefore, the three-class model was retained, as this model had comparable BIC (three-class model BIC = 6428.4; four-class model BIC = 6422.5), superior R^2^-entropy (three-class model R^2^-entropy = 0.81; four-class model R^2^-entropy = 0.77), and provided a stable solution. The three-class model yielded classes characterized by being “Healthy and wealthy males” (Class 1), “Aging, multimorbid, with socioeconomic burden” (Class 2), and “Young, healthy, and wealthy females” (Class 3) (Table 3 and Figure 1). Multimorbidity was present in 0%, 24%, and 4% of classes 1 to 3, respectively. The predominantly multimorbid class was characterized by being older with below average SEP and increased presence of risk factors.

**Table 3.** DA-dataset Latent Class Analysis conditional item response probabilities by variable per class in 671 Dutch adults from the general population. Data was collected between July 2024 and August 2024.

| Variable | Class 1 | Class 2 | Class 3 |
| --- | --- | --- | --- |
|  | *“Healthy and wealthy males”* | *“Aging, multimorbid, with socioeconomic burden”* | *“Young, healthy, and wealthy females”* |
| **Class Information** |  |  |  |
| Class population (N) | 189 | 325 | 157 |
| Estimated class proportion | 0.24 | 0.49 | 0.27 |
| Predicted by modal posterior | 0.28 | 0.48 | 0.23 |
| **Age** |  |  |  |
| 18 to 44 years | 0.48 | 0.00 | 0.91 |
| 45 years or older | 0.52 | 1.00 | 0.09 |
| **Sex** |  |  |  |
| Male | 1.00 | 0.53 | 0.10 |
| Female | 0.00 | 0.47 | 0.90 |
| **Socioeconomic Position** |  |  |  |
| Below average | 0.00 | 0.69 | 0.21 |
| Average | 0.08 | 0.15 | 0.31 |
| Above average | 0.92 | 0.16 | 0.48 |
| **Disease Status** |  |  |  |
| No chronic conditions | 0.79 | 0.39 | 0.82 |
| Single chronic condition | 0.21 | 0.37 | 0.14 |
| Multimorbidity | 0.00 | 0.24 | 0.04 |
| **Risk factors** |  |  |  |
| Absent | 0.69 | 0.42 | 0.79 |
| Present | 0.31 | 0.59 | 0.21 |
| **Depression** |  |  |  |
| Low depressive symptoms | 0.93 | 0.80 | 0.81 |
| Clinically relevant depressive symptoms | 0.07 | 0.20 | 0.19 |
| **Alexithymia** |  |  |  |
| No alexithymia | 0.87 | 0.73 | 0.77 |
| Alexithymia present | 0.13 | 0.28 | 0.23 |

**Figure 1.**
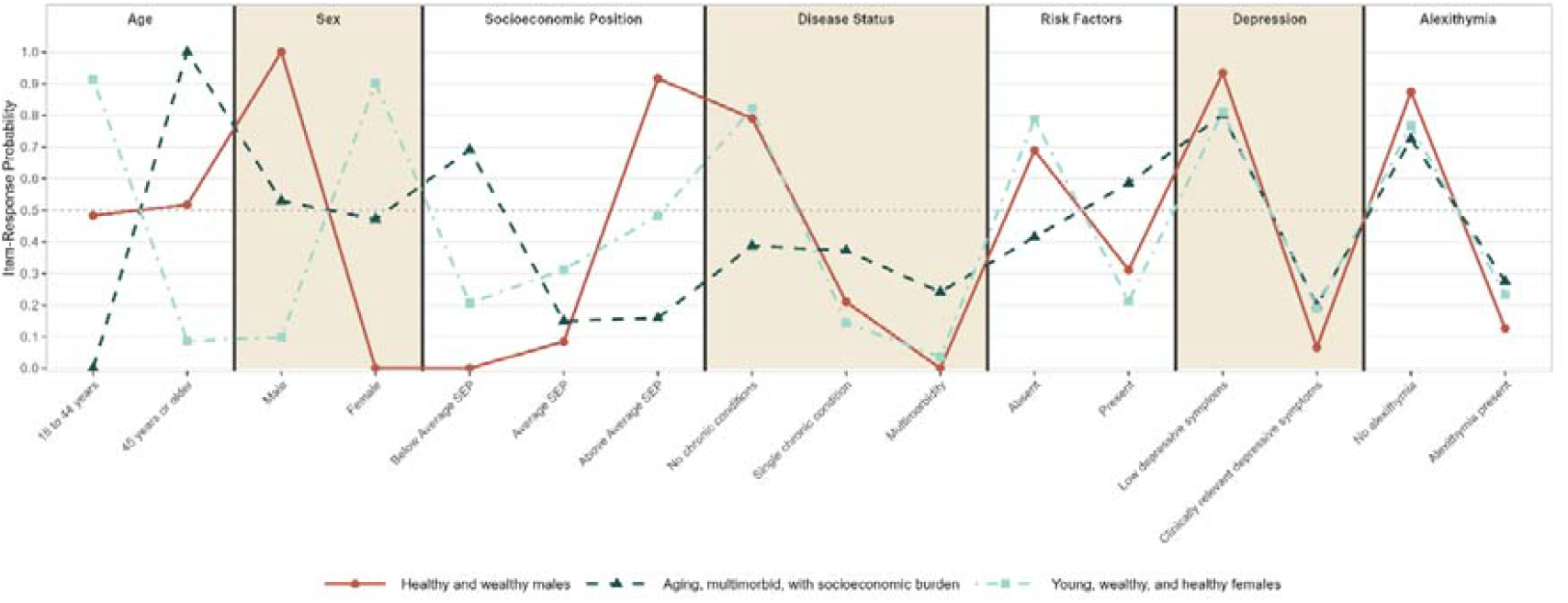
Visual representation of the DA-dataset Latent Class Analysis in 671 Dutch adults from the general population. Data was collected between July 2024 and August 2024.

In the L-dataset, a three-class model was found to be the best fit (BIC = 15361.0). The R^2^-entropy for this model was 0.65, indicating acceptable certainty of class separation. The three classes were indicative of being “Aging, multimorbid females with behavioral dysregulation” (Class 1), “Aging, healthy males with high self-regulation” (Class 2), and “Young, healthy, and wealthy” (Class 3) (Table 4 and Figure 2). The class members exhibited a multimorbidity prevalence of 22%, 12%, and 0%, respectively, in classes 1 to 3. In this dataset, the participants in the class with the highest multimorbidity prevalence were older, mostly female, and additionally exhibited below average SEP, increased presence of risk factors, and behavioral dysregulation (i.e., insufficient physical activity, below average self-regulation, and emotional eating).

**Table 4.** L-dataset Latent Class Analysis conditional item response probabilities by variable per class in 1022 Dutch adults from the general population. Data was collected between July 2024 and August 2024.

| Variable | Class 1 | Class 2 | Class 3 |
| --- | --- | --- | --- |
|  | *“Aging, multimorbid females with behavioral dysregulation”* | *“Aging, healthy males with high self-regulation”* | *“Young, healthy, and wealthy”* |
| **Class Information** |  |  |  |
| Class population (N) | 349 | 336 | 337 |
| Estimated class proportion | 0.35 | 0.32 | 0.34 |
| Predicted by modal posterior | 0.34 | 0.33 | 0.33 |
| **Age** |  |  |  |
| 18 to 44 years | 0.18 | 0.04 | 0.89 |
| 45 years or older | 0.82 | 0.96 | 0.11 |
| **Sex** |  |  |  |
| Male | 0.37 | 0.75 | 0.41 |
| Female | 0.63 | 0.25 | 0.60 |
| **Socioeconomic Position** |  |  |  |
| Below average | 0.48 | 0.38 | 0.15 |
| Average | 0.26 | 0.17 | 0.20 |
| Above average | 0.26 | 0.46 | 0.65 |
| **Disease Status** |  |  |  |
| No chronic conditions | 0.36 | 0.60 | 0.90 |
| Single chronic condition | 0.43 | 0.29 | 0.10 |
| Multimorbidity | 0.22 | 0.12 | 0.00 |
| **Risk factors** |  |  |  |
| Absent | 0.34 | 0.62 | 0.82 |
| Present | 0.66 | 0.39 | 0.18 |
| **Physical Activity** |  |  |  |
| Insufficiently active | 0.57 | 0.35 | 0.35 |
| Active | 0.43 | 0.65 | 0.65 |
| **Alcohol Use** |  |  |  |
| No likelihood | 0.89 | 0.88 | 0.83 |
| Likelihood present | 0.11 | 0.12 | 0.17 |
| **Smoking** |  |  |  |
| Non-smoker | 0.85 | 0.90 | 0.86 |
| Smoker | 0.15 | 0.10 | 0.14 |
| **Self-Regulation** |  |  |  |
| Below average self-regulation | 0.76 | 0.29 | 0.51 |
| Above average self-regulation | 0.24 | 0.71 | 0.49 |
| **Emotional Eating** |  |  |  |
| Non-emotional eater | 0.12 | 0.51 | 0.17 |
| Low emotional eater | 0.21 | 0.40 | 0.37 |
| Emotional to very emotional eater | 0.67 | 0.09 | 0.46 |
| **Sleep** |  |  |  |
| No disturbed sleep | 0.52 | 0.92 | 0.86 |
| Disturbed sleep | 0.48 | 0.08 | 0.14 |

**Figure 2.**
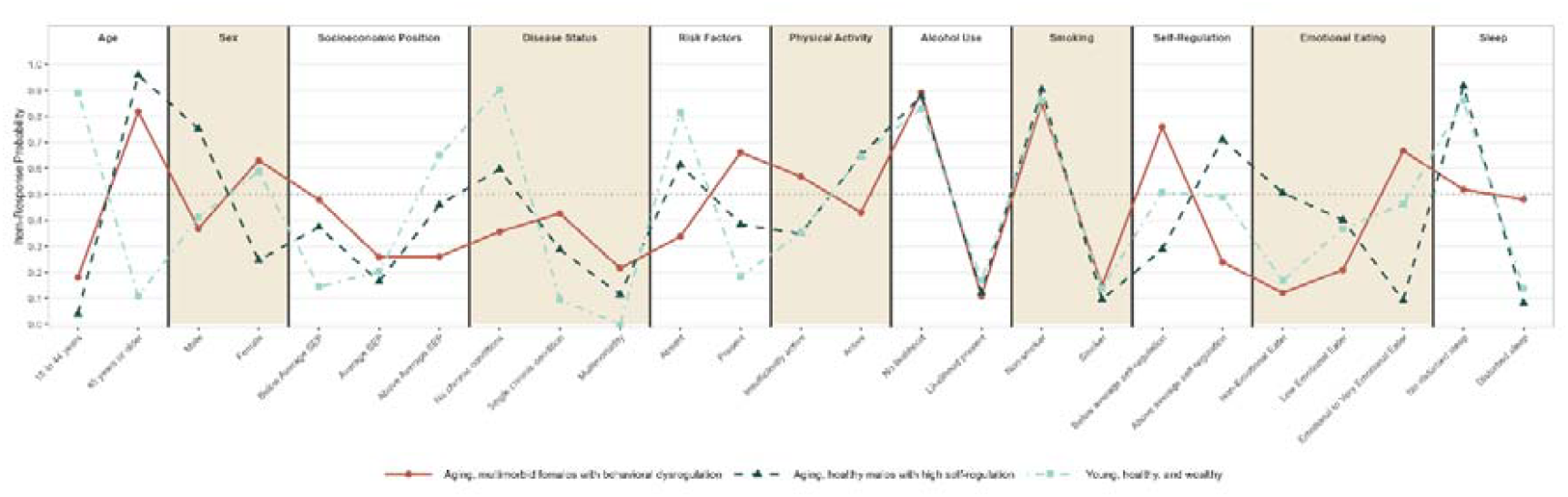
Visual representation of the L-dataset Latent Class Analysis in 1022 Dutch adults from the general population. Data was collected between July 2024 and August 2024.

### 3.3. Interoception

#### 3.3.1. Depression and Alexithymia Dataset

The regression analyses in the DA-dataset indicated that the multimorbid Class 2 had lower interoceptive accuracy compared to the healthier reference Class 1 (B = −1.14, 95% CI = [−2.89, 0.62]). Additionally, Class 2 had significantly higher interoceptive attention compared to Class 1 (B = 3.62, 95% CI = [0.97, 6.27]) (Table 5).

**Table 5.** DA-dataset regression analysis showing how latent classes predict interoceptive accuracy (IAS) and attention (IATS) scores in 671 Dutch adults from the general population. Data was collected between July 2024 and August 2024.

| Class | IAS (N = 671) |  |  |  | IATS (N = 671) |  |  |  |
| --- | --- | --- | --- | --- | --- | --- | --- | --- |
|  | B | SE | p | 95% CI | B | SE | p | 95% CI |
| Class 1 (reference) |  |  |  |  |  |  |  |  |
| Class 2 | -1.14 | 0.89 | 0.205 | [-2.89, 0.62] | 3.62 | 1.35 | 0.008* | [0.97, 6.27] |
| Class 3 | -3.31 | 1.06 | 0.002* | [-5.39, -1.24] | -0.20 | 1.59 | 0.898 | [-3.33, 2.93] |

IAS model statistics: $F(2,668) = 5.05$ , $p = 0.007$ , $AIC = 4969.3$ , $BIC = 4987.3$ , $RMSE = 9.78$ .
IATS model statistics: $F(2,668) = 5.31$ , $p = 0.005$ , $AIC = 5521.9$ , $BIC = 5539.9$ , $RMSE = 14.76$ .
Class 1: Healthy and wealthy males. Class 2: Aging, multimorbid, with socioeconomic burden. Class 3: Young, healthy, and wealthy females.

#### 3.3.2. Lifestyle Factors Dataset

L-dataset regression analyses showed that Class 1, characterized by multimorbid females with behavioral dysregulation, had significantly lower interoceptive accuracy compared to the healthier Class 2 (B = −2.36, 95% CI = [−3.83, −0.89]). Moreover, Class 1 had higher interoceptive attention in comparison to Class 2 (B = 1.07, 95% CI = [−1.42, 3.56]) (Table 6).

**Table 6.**
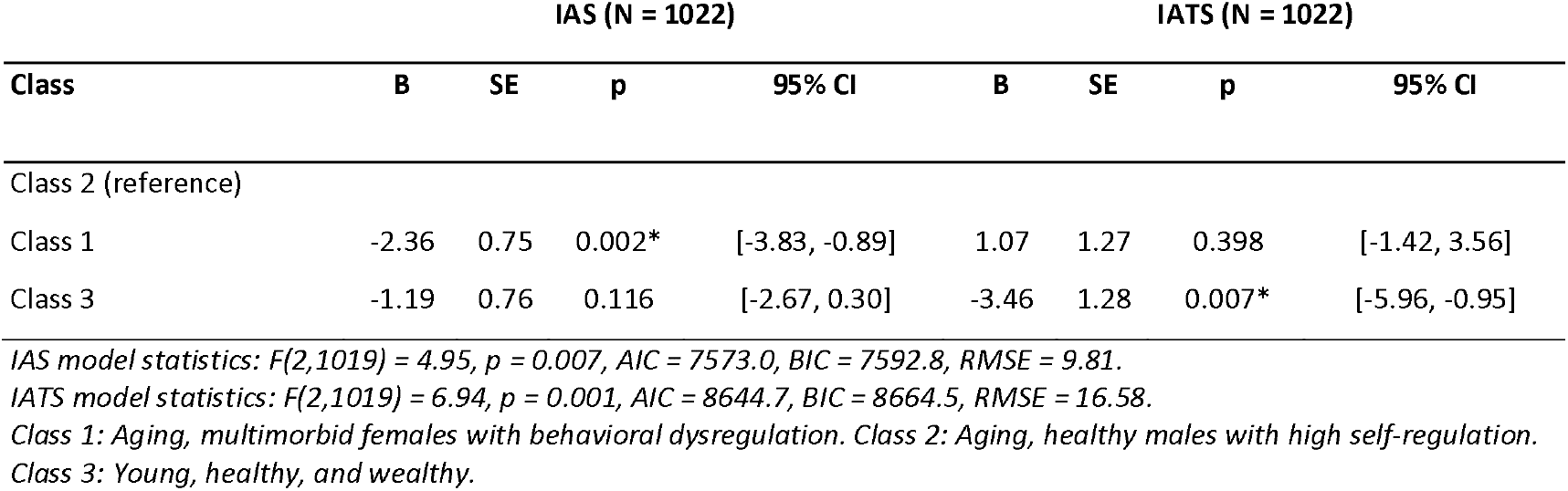
L-dataset regression analysis showing how latent classes predict interoceptive accuracy (IAS) and attention (IATS) scores in 1022 Dutch adults from the general population. Data was collected between July 2024 and August 2024.

#### 3.3.3. Sensitivity analysis

Sensitivity regression analyses in the DA-dataset, adjusted for age, sex, and depression, showed that the association between multimorbidity and interoceptive accuracy reversed direction (B = 1.04, 95% CI [−1.37, 3.46]). Similarly, the association between multimorbidity and interoceptive attention was attenuated to non-significance while maintaining its direction after adjustment (B = 2.19, 95% CI [−1.52, 5.89]). In the L-dataset, the association between multimorbidity and interoceptive accuracy remained significant and directionally consistent with the main analyses after adjusting for age and sex (B = −2.05, 95% CI [−3.62, −0.48]). The association between multimorbidity and interoceptive attention became statistically significant after adjustment (B = 3.23, 95% CI [0.60, 5.86]). Taken together, these findings suggest that associations between multimorbidity and interoceptive outcomes were suppressed or confounded by age, sex, and/or depressive symptoms, though the pattern was not consistent across datasets and outcomes.

## 4. Discussion

The current study aimed to examine how self-reported interoceptive accuracy and attention differ between subpopulations of Dutch adults with varying multimorbidity and psychosocial profiles.

Individuals characterized by multimorbidity, higher age, and lower SEP showed reduced interoceptive accuracy but increased interoceptive attention, suggesting heightened bodily awareness without accurate interpretation. These findings could indicate that multimorbid populations experience a distinct interoceptive profile characterized by hypervigilance to bodily signals accompanied by misinterpretation. This pattern may contribute to the psychosocial burden and self-management issues observed in multimorbid individuals, as maladaptive interoceptive processing may lead to adverse health behavior and symptom management.

While research specifically examining interoception in multimorbidity is limited, our findings that multimorbidity was associated with low interoceptive accuracy scores align with studies on interoception in people with chronic conditions, such as chronic pain and Parkinson’s disease (Locatelli et al., 2023). Similarly, reduced interoceptive accuracy is seen in individuals suffering from depression (Eggart et al., 2019; Quadt et al., 2018; Zhou et al., 2022). The multimorbid group in our study showed both higher rates of depressive symptoms and lower interoceptive accuracy compared to other groups. The link between depression and low interoceptive accuracy may indicate combined somatic and affective burden. Maladaptive interoceptive processing disrupts homeostasis by impairing regulation of internal bodily states, leading to stress responses, worsening physiological dysregulation, and complicated self-management in multimorbid individuals (Bayliss et al., 2007; Bonaz et al., 2021; Farb et al., 2015; Lee et al., 2024; Nord & Garfinkel, 2022). However, an important alternative explanation must be considered: depression is associated with negative self-evaluation and pessimistic view of one’s own abilities (Lou et al., 2019). As the IAS measures self-reported beliefs about interoceptive accuracy rather than objective interoceptive performance, lower IAS scores may partly reflect a generalized negative self-view rather than actual interoceptive impairment. Additionally, sensitivity analyses indicated depression, alongside age and sex, confounded interoceptive accuracy differences between the multimorbid and healthier classes. This suggests that rather than the presence of multiple chronic diseases alone multimorbid class membership covaries with older age, sex, and higher depressive symptom burden, each of which has been linked with altered interoceptive processing (Li et al., 2025; Prentice et al., 2022; Prentice & Murphy, 2022; Quadt et al., 2018). As depression appears to be a key driver of interoceptive differences between latent classes, multimorbid individuals may benefit from interventions targeting co-occurring interoceptive impairment and depressive symptoms. Together these findings point to interoceptive accuracy as a potentially important but understudied factor in the complex interplay between disease burden, affective symptoms, and self-management in multimorbidity.

In contrast, the current study identified increased interoceptive attention in multimorbid individuals. It is important to note that increased interoceptive attention should be interpreted in conjunction with interoceptive accuracy, as heightened attention to bodily signals can be either adaptive or maladaptive depending on the degree to which these signals are perceived accurately. This hypervigilance towards bodily signals despite low interoceptive accuracy may reflect a compensatory but maladaptive response. A similar pattern is observed in anxiety disorders: individuals at risk of anxiety struggle to distinguish between potentially aversive bodily signals and constantly ongoing and fluctuating interoceptive signals (Bonaz et al., 2021; Paulus & Stein, 2010; Quadt et al., 2018). Consequently, these signals are accompanied by negative emotions, resulting in worrying through increased production of thoughts and associated beliefs. This pattern, where interoceptive signals are misinterpreted as threatening due to negative predictions, may also be present in multimorbid individuals, as their disease status heightens health-related concerns.

These interoceptive patterns may be linked to the broader behavioral vulnerabilities observed in multimorbid individuals, including low physical activity, poor self-regulation, emotional eating, increased smoking, and more sleep disturbances. Interoception likely plays an important role in the relationship between multimorbidity and lifestyle behaviors (Mulder et al., 2025). For example, low physical activity is known to be associated with multimorbidity (Delpino et al., 2022), while regular physical activity may improve interoceptive abilities over time (Amaya et al., 2021). The current findings showed that physical inactivity in multimorbid individuals was associated with increased interoceptive attention but decreased accuracy. This could be because of a vicious cycle: low self-regulation leads to less physical activity participation (Ylitalo et al., 2023), and being less physically active leads to poor interoceptive accuracy, resulting in the protective role of physical activity in disease development not being fully utilized (Dhalwani et al., 2016). This illustrates the importance of the associations between interoception and behavioral regulation.

The relationship between interoception and substance use in this sample was complex. Surprisingly, alcohol dependence did not differ substantially between multimorbid and healthier groups, despite previous findings linking impaired interoception to higher alcohol use (Wisniewski et al., 2021). This may reflect age-related reductions in drinking among chronically ill individuals, as the multimorbid group was also older in age compared to at least one other class (Alvarez-Galvez et al., 2023). Alternatively, the alcohol-harm paradox may be an explanation: this paradox states that socioeconomically disadvantaged groups experience greater harm due to alcohol consumption, despite comparable or even lower alcohol intake (Katikireddi et al., 2017). Thus, the slightly lower likelihood of alcohol dependence observed in the multimorbid group does not necessarily indicate a lower alcohol-related health burden, but may contribute disproportionately to adverse health outcomes in an already vulnerable population. Additionally, lower alcohol dependence does not necessarily reflect healthier behavior, but may coexist with other maladaptive coping strategies. The multimorbid group showed more physical inactivity, below average self-regulation, higher rates of emotional eating, and elevated sleep disturbances, suggesting a shift toward other maladaptive behaviors rather than overall improved lifestyle patterns.

When interpreting these findings, both methodological strengths and limitations must be considered. A key strength of the current study was the use of latent class analyses to identify distinct profiles of multimorbid subgroups within a sample representative of the general Dutch population, allowing for the clustering of multiple co-occurring conditions and strengthening generalizability to the broader Dutch adult population. In addition, two domains of interoception (i.e., accuracy and attention) were examined, providing a complete picture of the complex relation between multimorbidity and interoception.

Several limitations must be acknowledged. First, dichotomizing continuous and ordinal variables may have reduced variability and obscured nuance in class differentiation. This also relates to a second limitation, namely that the nature of the datasets imposed constraints. In one dataset, depression categories had to be collapsed due to low frequencies. In the second dataset the same had to be done for emotional eating categories. Collapsing categories may have led to loss of information and less nuanced differences between classes. Second, it must be acknowledged that latent class analyses are sensitive to sample size. While the current sample size exceeded thresholds suggested by literature (Weller et al., 2020), and fit indices supported our class solutions, it may be that our findings are not replicable in samples with different sizes. Third, labeling of classes requires some degree of subjectivity. While we have labelled the classes in a way that we believe captures their most distinctive features, these labels are a simplification. Thus, in clinical applications, labels should be treated as simplistic communication of class profiles rather than diagnostic categories. Finally, it needs to be mentioned that in the current study only self-reported data was used, which may cause potential misclassification and information bias. Despite these limitations, the current study offers valuable insight into the role of interoception in multimorbidity within a real-world adult population.

These findings have important implications for both research and clinical practice. This study identified distinct multimorbid subgroups in Dutch adults characterized by psychosocial vulnerability and maladaptive interoceptive processes, highlighting the complex interplay between physical illness, mental health, and behavioral regulation. This complexity is further demonstrated by our sensitivity analyses: associations between multimorbidity and interoception showed varying patterns across datasets and interoceptive dimensions, with the associations between multimorbidity and interoceptive accuracy in the DA-dataset even changing direction compared to the main analysis. This may mean that age, sex, and/or depression function as mediators in the causal pathway between multimorbidity and interoceptive accuracy, resulting in over-adjustment when controlling for these variables (Schisterman et al., 2009). Alternatively, these covariates may act as colliders: age, sex, and/or depression may be influenced by both multimorbidity and interoceptive accuracy, and controlling for them induces deceptive associations (Greenland, 2003). Additionally, recent evidence suggests that interoceptive domains (e.g., accuracy or attention) and modalities (e.g., heartrate, respiratory, or gastrointestinal) associate differently with various diseases (Schoeller et al., 2025). The current study demonstrated such differences between interoceptive accuracy and attention in multimorbid individuals. Future research should also examine whether interoceptive profiles of multimorbid individuals differ between modalities, to support the development of more targeted, personalized intervention programs.

Although interoception-informed interventions show promise, evidence in complex multimorbid populations remains scarce (Smith et al., 2021). Multimorbid patients face psychosocial burdens and often experience maladaptive interoception, which may impair treatment adherence, self-management, and help-seeking behavior. This calls for a shift toward patient-centered and integrated care that accounts for patients’ biological, psychological, and social contexts. We propose that embedding interoceptive assessment in multimorbid care may facilitate the continuous adjustment of treatment strategies. By identifying psychosocial burdens alongside physical symptoms, care delivery can be better aligned with patients’ evolving needs.

Interventions that enhance bodily awareness offer additional opportunities to support patients: for instance, mindfulness, breathing techniques, yoga, and the Feldenkrais method have shown potential to improve interoception and emotional regulation (Berland et al., 2022; Fissler et al., 2016; Nord & Garfinkel, 2022; Weng et al., 2021). Implementing such interventions within multimorbidity care may strengthen patients’ adherence to interventions or treatments and improve self-management. Strengthening interoceptive abilities in patients may not only improve treatment efficacy, but also improve patient engagement in integrated care.

## 5. Conclusion

This study highlights the psychosocial complexity of Dutch multimorbid populations which may impact their self-management and health behavior. Notably, the multimorbid population demonstrated reduced interoceptive accuracy alongside increased attention that may further exacerbate their health and psychosocial challenges. The findings underscore the need to expand treatments to include psychosocial domains for multimorbid patients. This work provides a foundation for future research and intervention development, marking an important step toward interoception-informed care for multimorbid populations.

## 6. Declarations

### Declaration of competing interests

The authors declare there are no conflicts of interest.

### Funding statement

This study is part of a research project that is funded by the Velux Stiftung (project number 1815). The sponsor had no influence on the content of this article.

### Author contributions

JM: Conceptualization, Methodology, Formal analysis, Investigation, Data Curation, Writing – Original Draft, Writing – Review & Editing, Visualization, Supervision, Project Administration; CMB: Data Curation, Methodology, Formal Analysis, Writing – Original Draft, Writing – Review & Editing, Visualization; AKS: Writing – Review & Editing; JCK: Conceptualization, Methodology, Writing – Review & Editing, Supervision, Funding Acquisition.

### Data availability

Datasets will be made available by the corresponding author upon reasonable request. Analysis scripts are available on the following repository: https://github.com/JesperMulder/interoception-multimorbidity.

### Informed consent

Informed consent was obtained from all individuals participating in this study.

### Declaration of Generative AI and AI-Assisted Technologies in the Writing Process

During the preparation of this work the author(s) used a large language model in order to help with writing the code for statistical analyses. After using this tool/service, the author(s) reviewed and edited the content as needed and take(s) full responsibility for the content of the published article.

## Notes

### Competing Interest Statement

The authors have declared no competing interest.

### Author Declarations

The research committee of the Leiden University Medical Center Department of Public Health and Primary Care (reference number WSC-2024-26/ SP) and medical ethical committee Leiden-The Hague-Delft (METC number N24.072) approved this study as non-WMO.

